# Multidimensional Evaluation of Large Language Models on the AAP In-Service Examination: Assessing Accuracy, Calibration, and Citation Reliability

**DOI:** 10.1101/2025.10.14.25338040

**Authors:** Prita Abhay Dhaimade, Robin Henderson

## Abstract

**Background:** Large language models (LLMs) have demonstrated rapid advancements in natural language understanding and generation, prompting their integration into biomedical research, clinical practice, and professional education. However, systematic evaluation of LLMs in specialty-specific domains such as dentistry and periodontology remain limited, particularly regarding multidimensional performance metrics.

**Objective:** To conduct a comprehensive, multidimensional assessment of commercially available LLMs: GPT-4.0, GPT-5.0, and Claude SONNET 4.0 on the American Academy of Periodontology in-service examination, focusing on response accuracy, self-assessed confidence calibration, citation validity, and hallucination prevalence.

**Methods:** Models were evaluated on the 2024 AAP In-Service Examination (331 questions) using two formats: Full Test (all questions at once) and Individual Question (one at a time). Prompts were standardized; models selected answers, and for GPT-5.0 and Claude SONNET 4.0, also provided confidence ratings and citations. Citation validity was assessed using a human-in-the-loop protocol with expert review. Statistical analyses included chi-square, McNemar’s, and logistic regression to assess accuracy, question fatigue, confidence calibration, and citation reliability.

**Results:** LLMs achieved high overall accuracy (78–87%), with the Individual Question format consistently yielding higher scores than Full Test, though differences were not statistically significant.

Accuracy was highest in fact-dense domains (biochemistry, physiology, microbiology) and lowest in integrative domains (diagnosis, therapy). Significant question fatigue was observed in GPT-5.0 Full Test mode (OR = 0.997, p = 0.035), but not in Individual Question mode.

Confidence scores predicted accuracy, with the strongest calibration in Individual Question mode. Citation analysis revealed frequent hallucinations, mostly critically erroneous, and citation validity was independent of answer accuracy.

**Conclusions:** LLMs can answer a broad spectrum of periodontal specialty questions, but their reliability varies with context and information presentation. While promising as adjunctive tools, their outputs— especially for complex reasoning and citations—require rigorous human review in educational and research settings to ensure accuracy and safety.

**Author Summary:** Artificial intelligence chatbots are rapidly entering medical education, yet we lack comprehensive understanding of their reliability when students depend on them for learning. We developed a multidimensional evaluation framework to systematically assess AI performance beyond simple accuracy, examining how these systems behave across different medical topics, question types, and presentation formats.

Using 331 real dental examination questions, we tested three major AI systems, analyzing not only correctness but also confidence calibration - whether AI confidence levels match actual accuracy - and implementing human-in-the-loop verification to check if cited sources actually exist.

Our findings highlight critical vulnerabilities in current AI systems. Most alarmingly, these chatbots fabricated nearly half of their citations while maintaining unwavering confidence in both correct and incorrect responses. This combination of overconfidence and misinformation means students cannot distinguish reliable from unreliable AI responses. Additionally, we documented progressive performance decline during sequential questioning, similar to human cognitive fatigue.

While we know AI systems generate rather than retrieve information, our research demonstrates the real-world consequences of this limitation. As artificial intelligence integrates into education, healthcare diagnostics, and insurance decisions, these findings underscore the urgent need for better evaluation frameworks and user education about AI limitations.

## Introduction

Artificial intelligence (AI) has rapidly evolved into a transformative technology across scientific and clinical disciplines. Among its most significant developments are large language models (LLMs), which are trained on vast corpora of text to generate coherent, human-like language. Their ability to recognize linguistic patterns and predict plausible continuations has enabled widespread application in knowledge retrieval, summarization, translation, and dialogue systems. Their integration into healthcare has been particularly notable, with applications ranging from clinical documentation and literature synthesis to diagnostic support and educational innovation.

Dentistry has begun to embrace AI in parallel with medicine, with early work demonstrating utility in diagnostic imaging, caries detection, treatment planning, and predictive analytics. (1) More recently, LLMs have been explored as tools for dental education and clinical training, where they may supplement student learning, provide rapid access to foundational knowledge, and support preparation for standardized examinations. (2,3)

It is important to recognize, however, that LLMs are not designed to discover or verify truth. Instead, they function as statistical models that predict the most probable sequence of words given prior input. This capability allows them to generate fluent and contextually appropriate language, but it also constrains their factual reliability. Unlike structured databases, LLMs do not store or retrieve information in a queryable format. Although their outputs can be supplemented by external databases or curated sources, current methods provide limited transparency or control over how such sources are integrated or weighted. (4)

These limitations are especially concerning in domains where factual precision is critical, such as medicine and research. Consequently, the systematic evaluation of LLMs has become a central priority, with growing efforts devoted to developing robust metrics and standardized approaches for assessing their performance across multiple dimensions.

To date, much of the research evaluating LLMs’ performance and accuracy in medicine has employed standardized, high-stakes assessments such as the United States Medical Licensing Examination (USMLE), with systematic reviews confirming variable but generally improving performance across model generations. (5–7)In dentistry, similar efforts have examined LLM performance on the National Board Dental Examination (NBDE/INBDE) and related professional assessments(8–10), while a recent study in the *Journal of Periodontology* reported on the use of the AAP Periodontics In-Service Examination to benchmark ChatGPT against specialty-level questions(11). These studies underscore the value of standardized examinations as rigorous testbeds for evaluating LLM performance. While such studies primarily assess accuracy and reasoning, they have largely overlooked the ability of LLMs to provide verifiable citations. The widespread use of these models has revealed a consistent problem of fabricated or “hallucinated” references, where outputs may include plausible sounding but nonexistent sources or misattributed citations. (12)This issue is particularly problematic in educational settings, where students may lack the experience to critically evaluate references and risk accepting fabricated citations as valid. Such limitations not only undermine trust in AI-assisted learning but also pose risks to the integrity of scholarly work.

Given these gaps, the present study was designed to provide a multidimensional evaluation of LLM performance in periodontology using the AAP In-Service Examination. The objectives were to compare model accuracy across successive generations, assess the influence of prompt optimization and test presentation format on performance, quantify the rate of citation hallucination, and evaluate the validity of self-reported confidence in responses. By employing a specialty-specific professional examination as the test environment, this study aimed to generate a comprehensive assessment of LLM capabilities and limitations in a context where factual reliability, citation integrity, and calibrated confidence are critical.

## Methods

### Study Design

This study employed a comparative performance assessment framework to evaluate large language models (LLMs) in the domain of periodontology. A controlled experimental design (Figure 1) was used to ensure reproducibility and transparency. Evaluation focused on two dimensions, response accuracy and a human-in-the-loop (HITL) protocol to validate the reliability of model-generated citations. Three transformer-based models were selected based on published performance benchmarks and multimodal capabilities: ChatGPT-4.0 and ChatGPT-5.0 (OpenAI, San Francisco, CA, USA), and Claude SONNET 4.0 (Anthropic, San Francisco, CA, USA). The citation validity was tested only for Chat GPT 5.0 (OpenAI, San Francisco, CA, USA), and Claude SONNET 4.0 (Anthropic, San Francisco, CA, USA).

**Figure 1:**
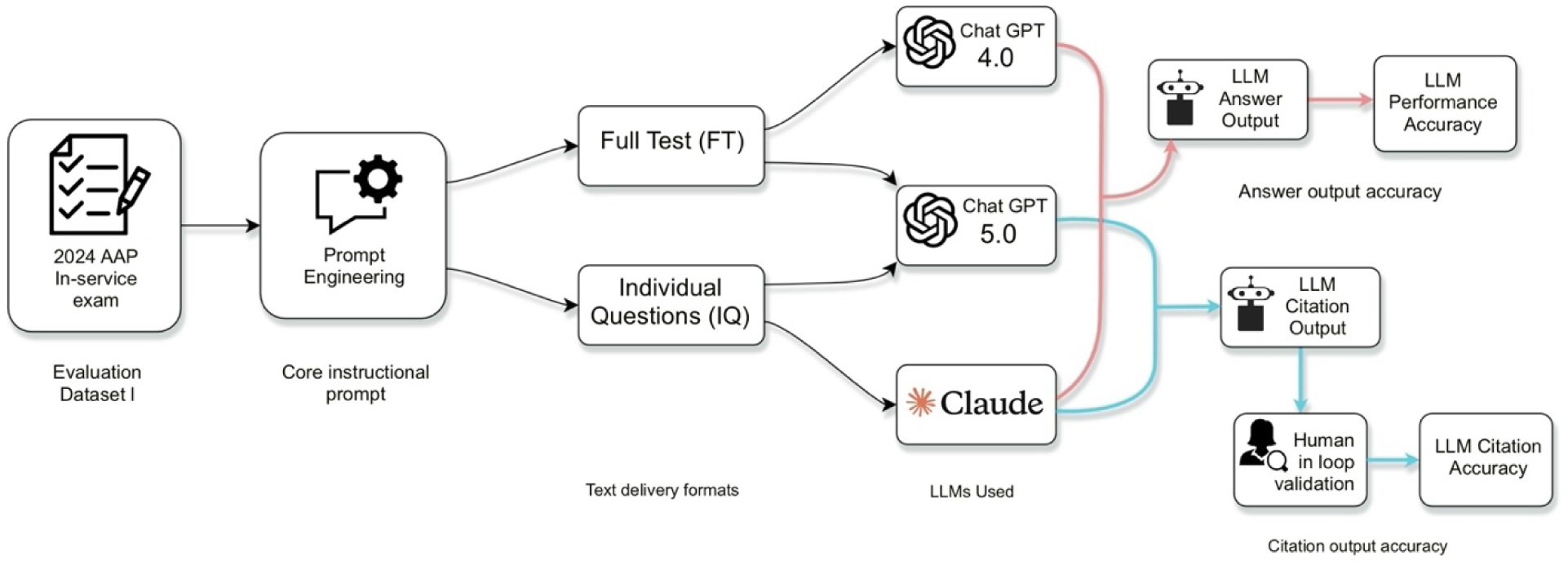
Study flow for evaluating large language model performance on a specialty periodontology examination

### Evaluation Dataset

The 2024 American Academy of Periodontology (AAP) In-Service Examination, comprising 331 multiple-choice questions, was used as the evaluation instrument. For structured analysis, the investigators categorized the items on the exam into four domains: general factual knowledge, author-specific/ study specifics factual knowledge, analytical reasoning, and image-based question.

### Prompt Design

Two exam delivery formats were employed. In the Full Test (FT) format, the entire question set was presented simultaneously, simulating unrestricted access. In the Individual Question (IQ) format, questions were delivered one at a time, mirroring the experience of a traditional test. Both ChatGPT models were evaluated under FT and IQ conditions, whereas Claude SONNET 4.0 was assessed only in IQ format.

Prompt formulation was standardized across all experimental conditions to minimize variability and ensure that observed differences in performance reflected intrinsic model capabilities rather than disparities in instruction. All models were required to select the most appropriate answer choice for each multiple-choice question, and prompts were written to be unambiguous, concise, and task-specific, with contextual information included only when necessary to support question comprehension. A single core instruction was employed for baseline evaluation: <u>“Select the single most accurate answer choice for the following multiple-choice question using all relevant knowledge and sources available to you.”</u> For ChatGPT-5.0 and Claude SONNET 4.0, additional instructions were included to (1) indicate their confidence in the selected answer as a percentage (0–100%), and (2) provide verifiable citations including Digital Object Identifiers (DOIs), PubMed identifiers (PMIDs), or persistent URLs to support their response choice. These additional tasks were applied exclusively to these models to enable assessment of confidence calibration and citation validity alongside overall accuracy.

### Human-in-the-Loop Citation Validation

A human-in-the-loop protocol was implemented specifically for citation validation.

References generated by ChatGPT-5.0 and Claude SONNET 4.0 were reviewed by a single evaluator trained in biomedical literature retrieval. Each citation was categorized using a three-tier taxonomy: Completely valid-existing and accessible citations accurately supporting the information provided, partially correct – real citations with minor inaccuracies or partial claim support, critically wrong – fabricated, inaccessible, or unrelated citations. This process enabled systematic detection of hallucinated or misattributed references.

## Statistical Analysis

Statistical analyses were conducted using IBM SPSS Statistics (version 31, IBM Corp., Armonk, NY, USA) and Python (version 3.12.10) to evaluate model performance, assess citation validity, and explore secondary relationships. Descriptive statistics and chi-square tests of independence were used to summarize and compare response accuracy across models.

Within each model version, the effect of question presentation format was evaluated using chi-square tests, and McNemar’s test was applied to discordant question-level pairs. For models that generated confidence scores (GPT-5.0 and Claude SONNET 4.0), logistic regression was used to examine the relationship between confidence and accuracy. Citation validity was analyzed by categorizing references into predefined validity groups and comparing their distributions between models using chi-square tests. The potential effect of token length on accuracy was investigated using question position as a proxy, with logistic regression modeling accuracy as the dependent variable and question index as a continuous predictor, and the Cochran–Armitage trend test applied to assess linear trends across ordered question bins.

## Results

### Overall Model Performance

A total of 331 multiple-choice questions from the 2024 AAP In-Service Examination were evaluated across four experimental conditions: GPT-4.0 Full Test (FT), GPT-4.0 Individual Question (IQ), GPT-5.0 FT, and GPT-5.0 IQ. Overall model accuracy varied significantly by version and delivery format. GPT-5.0 achieved the highest performance in IQ mode, correctly answering 279 of 331 questions (84.29%), compared with 274 of 331 (82.78%) for GPT-4.0 IQ. However, contrary to the individual question results, GPT-5.0 in FT mode achieved 80.85% accuracy (266 of 329 questions), substantially higher than GPT-4.0 FT at 78.12% (257 of 329 questions).

### Section and domain specific performance patterns

A network visualization of model performance across the 2024 American Academy of Periodontology In-Service Examination demonstrated consistent topic-dependent trends (Figure 2). The visualization illustrates the distribution of model accuracy across nine content topics, with nodes representing large language model configurations (blue) and examination topics (yellow), and edge color and thickness indicating performance levels (green: ≥0.90, red: 0.80– 0.89, gray: <0.80). Descriptive analysis revealed that certain subject areas exhibited systematically higher accuracy scores across all tested models, particularly Biochemistry and Physiology (mean accuracy = 0.968) and Microbiology and Immunology (mean accuracy = 0.960), which can also be visualized as green connections (≥0.9 accuracy) with multiple LLMs in the network visualization. Conversely, several domains proved universally challenging, with Diagnosis (mean accuracy = 0.710) and Periodontal Therapy (mean accuracy = 0.748) showing predominantly red connections (<0.8 accuracy) across all model variants.

**Figure 2:**
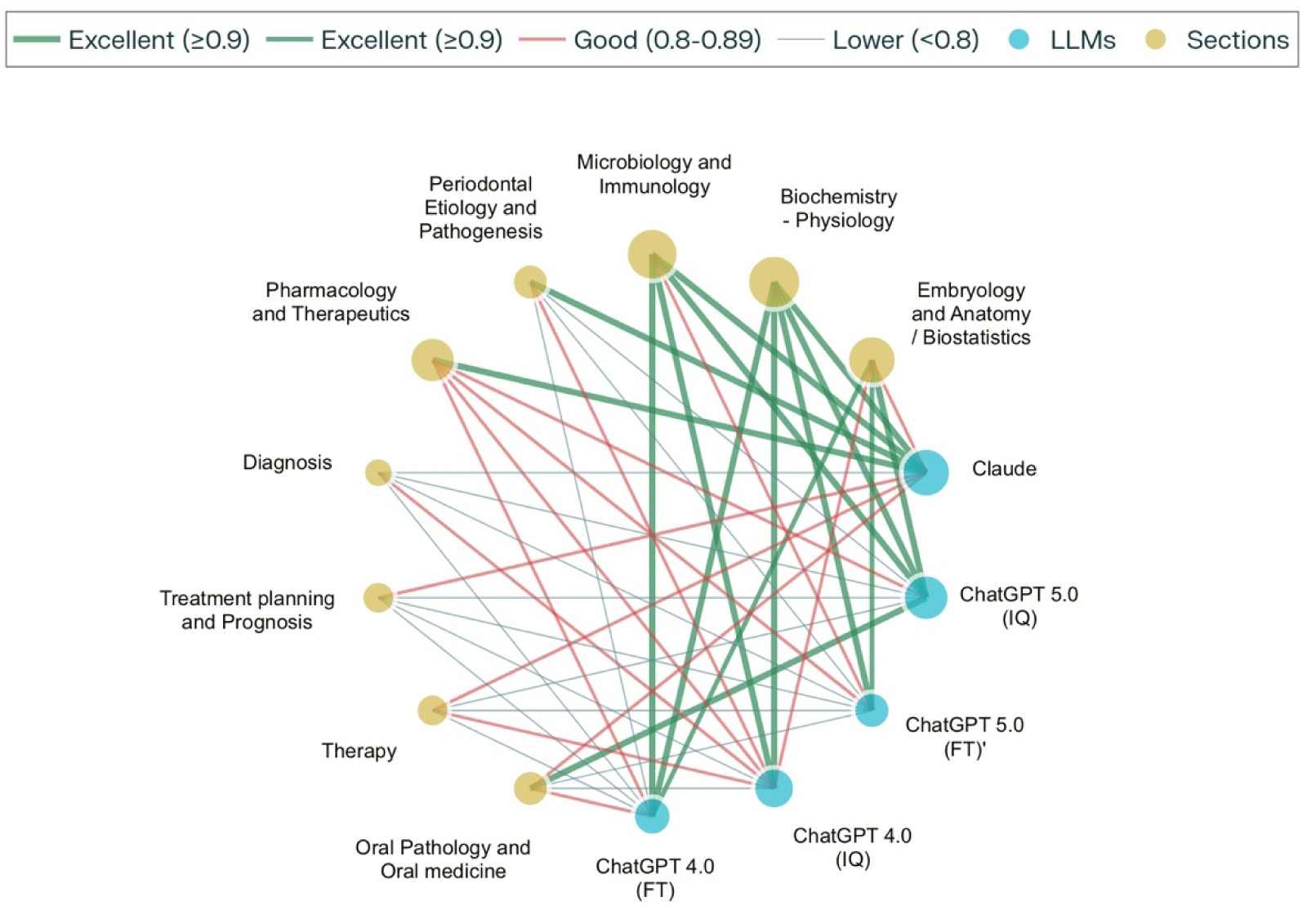
Network visualization of large language model performance across examination sections.

This pattern remained consistent despite variations in model training approaches, with both Full test and individual question versions of ChatGPT 4.0 and 5.0, as well as Claude SONNET 4.0, exhibiting similar relative performance hierarchies across subject domains. The network visualization further reveals that while individual models varied in overall performance— with Claude SONNET 4.0 achieving the highest overall accuracy (0.874) and ChatGPT 5.0FT the lowest (0.799)—the ranking of subject difficulty remained remarkably stable, suggesting that certain dental education domains present inherent challenges for current large language model architectures that transcend specific training methodologies or model parameters.

Figure 3 demonstrates marked heterogeneity in large language model (LLM) performance across dental education question taxonomies, revealing distinct competency profiles that transcend model architecture and training methodologies. Radar chart analysis of 331 questions across four cognitive domains—factual recall (n=186), author-referenced queries (n=126), image-based interpretation (n=10), and analytical reasoning (n=9)—exposed systematic performance variations that remained consistent across five state-of-the-art models: ChatGPT 4.0 (FT & IQ), ChatGPT 5.0 (FT &IQ), and Claude SONNET 4.0.

**Figure 3:**
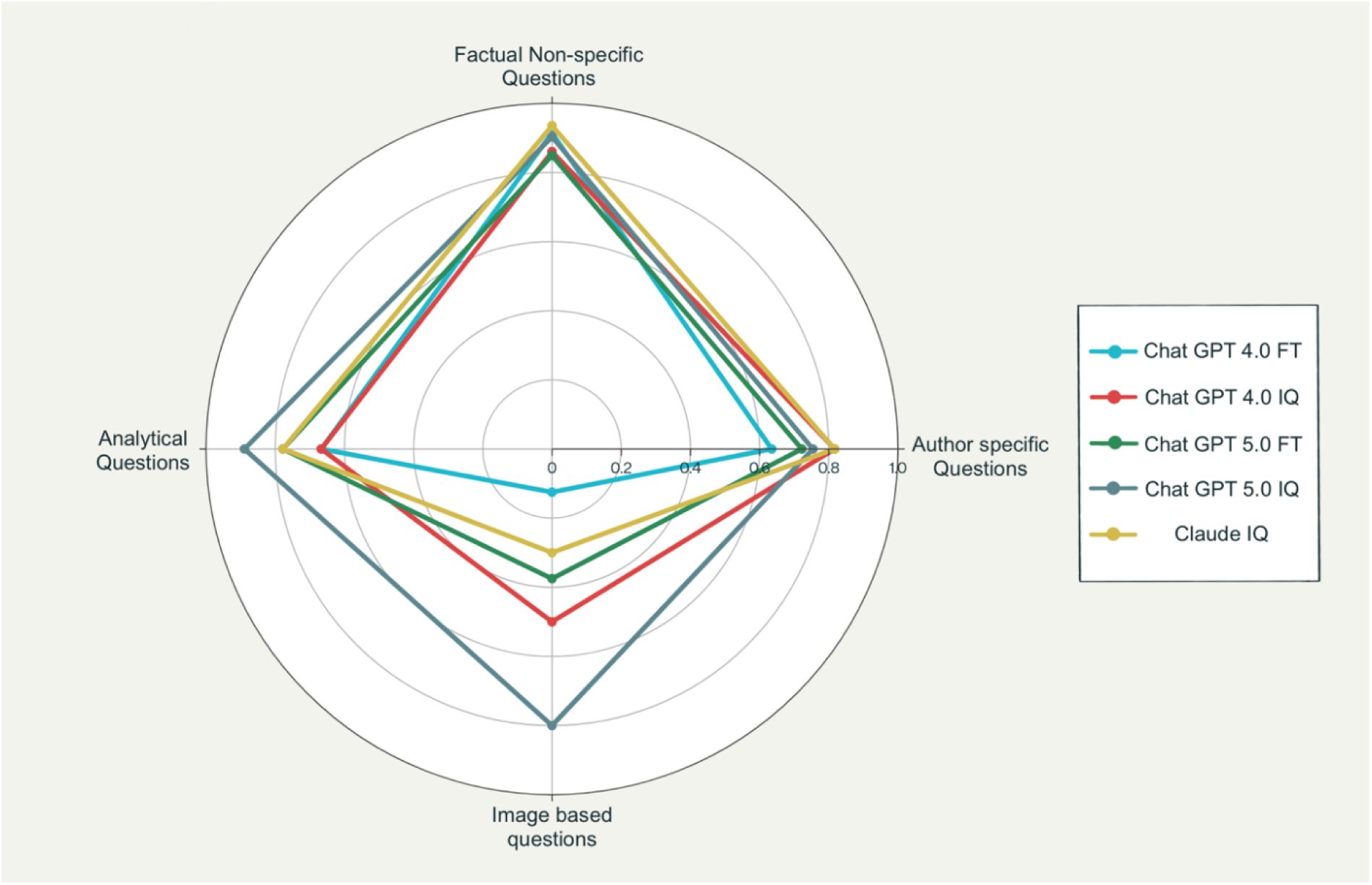
Radar plot comparing model accuracy across four question categories—factual, author-specific, analytical, and image-based—for all LLM configurations.

### Comparison within successive generations of LLMs: Chat GPT 4.0 vs. Chat GPT 5.0

A chi-square test of independence was conducted to compare the overall accuracy of GPT-4.0 and GPT-5.0 under Full Test (FT), Individual Question (IQ), and combined conditions. GPT-5.0 exhibited modest but consistent improvements in accuracy across all testing conditions, although none of the differences reached statistical significance (α = 0.05) (Table 4). The largest difference was observed in the FT condition (+2.74 percentage points; χ² = 0.596, *p* = 0.440), while the improvement in the IQ condition was smaller (+1.51 percentage points; χ² = 0.176, *p* = 0.675). Overall, GPT-5.0 achieved 82.58% accuracy compared with 80.45% for GPT-4.0 (χ² = 0.850, *p* = 0.357), and all effect sizes indicated small magnitudes of difference (Cramer’s V ≤ 0.030). These findings suggest incremental advancements in model capability between generations, though without statistically significant performance gains.

### Effect of Presentation Format (Full Test (FT) vs. Individual Question (IQ))

Comparison of model performance across presentation formats revealed a consistent pattern favoring the Individual Question (IQ) condition over the Full Test (FT) condition (Table 2). Although differences did not reach conventional levels of statistical significance (α = 0.05), IQ presentation was associated with higher accuracy across all analyses. GPT-4.0 achieved 82.98% accuracy in IQ format compared to 78.12% in FT format, representing a 4.86 percentage point improvement (χ² = 2.18, *p* = 0.140). GPT-5.0 demonstrated a similar directional effect, with accuracy improving from 80.85% (FT) to 84.19% (IQ), a 3.34 percentage point increase (χ² = 1.05, *p* = 0.305). When both models were combined, overall performance improved by 4.10 percentage points (χ² = 3.41, *p* = 0.065), approaching statistical significance (Cramer’s V = 0.051).

**Table 1:** Descriptive performance of large language models (LLMs) across subject domains and testing conditions. Accuracy of GPT-4.0 and GPT-5.0 was evaluated using the 2024 American Academy of Periodontology (AAP) In-Service Examination (331 multiple-choice questions). Results are presented as the number and percentage of correct responses for each section under Full Test (FT) and Individual Question (IQ) conditions. Completion rates and missing responses are also reported. IQ presentation consistently yielded higher accuracy than FT for both models, and GPT-5.0 outperformed GPT-4.0 across all sections.

| Section | GPT-4.0 FT | GPT-4.0 IQ | GPT-5.0 FT | GPT-5.0 IQ | Claude SONNET 4.0 |
| --- | --- | --- | --- | --- | --- |
| Embryology and Anatomy / Biostatistics | 25/29 (86.21%) | 26/29 (89.66%) | 25/29 (86.21%) | 27/29 (93.10%) | 26/29 (89.66%) |
| Biochemistry-Physiology | 29/31 (93.55%) | 30/31 (96.77%) | 31/31 (100.00%) | 29/31 (93.55%) | 31/31 (100.00%) |
| Microbiology & immunology | 25/25 (100.00%) | 25/25 (100.00%) | 22/25 (88.00%) | 24/25 (96.00%) | 24/25 (96.00%) |
| Periodontal Etiology and Pathogenesis | 28/37 (75.68%) | 30/37 (81.08%) | 26/37 (70.27%) | 29/37 (78.38%) | 34/37 (91.89%) |
| Pharmacology & Therapeutics | 32/36 (88.89%) | 31/36 (86.11%) | 29/36 (80.56%) | 32/36 (88.89%) | 35/36 (97.22%) |
| Diagnosis | 23/29 (79.31%) | 24/29 (82.76%) | 19/29 (65.52%) | 19/29 (65.52%) | 18/29 (62.07%) |
| Treatment planning and Prognosis | 19/27 (70.37%) | 20/27 (74.07%) | 20/27 (74.07%) | 21/27 (77.78%) | 22/27 (81.48%) |
| Therapy | 42/77 (54.55%) | 65/77 (84.42%) | 58/77 (75.32%) | 61/77 (79.22%) | 62/77 (80.52%) |
| Oral Pathology / Oral Medicine | 34/40 (85.00%) | 23/40 (57.50%) | 29/40 (72.50%) | 37/40 (92.50%) | 35/40 (87.50%) |
| Total | 257/331 (77.64%) | 274/331 (82.78%) | 259/331 (78.25%) | 279/331 (84.29%) | 287/331 (86.71%) |

**Table 2:**
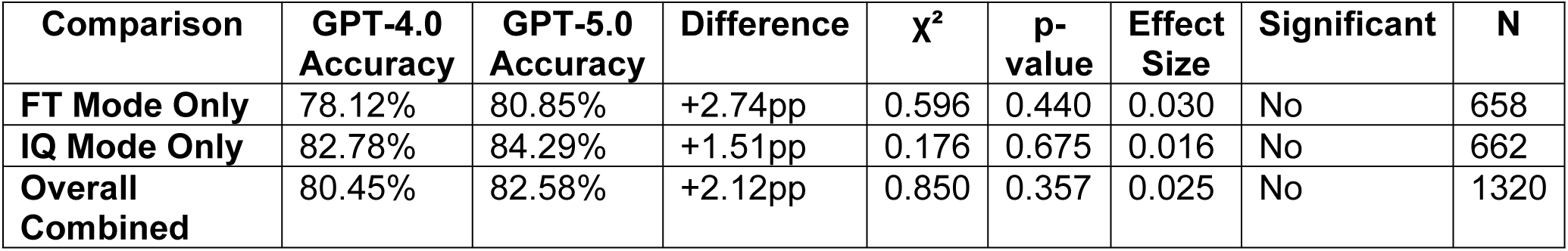
Chi-square test of independence comparing GPT-4.0 and GPT-5.0 performance across presentation conditions.

**Table 3:**
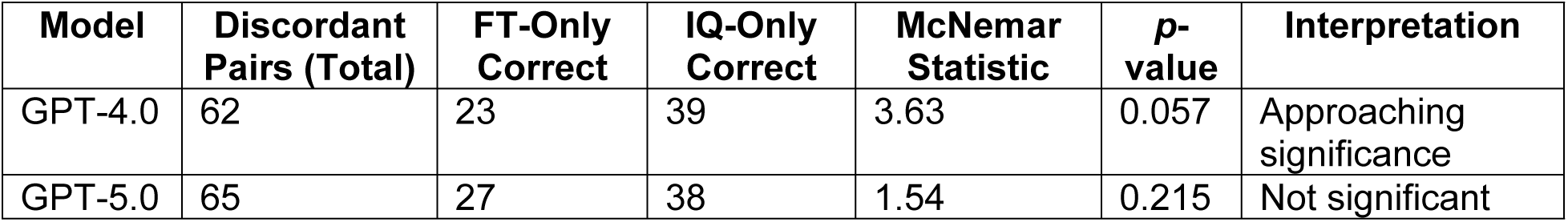
McNemar’s test results for paired comparison of Full Test and Individual Question conditions.

| Model | Discordant Pairs (Total) | FT-Only Correct | IQ-Only Correct | McNemar Statistic | p-value | Interpretation |
| --- | --- | --- | --- | --- | --- | --- |
| GPT-4.0 | 62 | 23 | 39 | 3.63 | 0.057 | Approaching significance |
| GPT-5.0 | 65 | 27 | 38 | 1.54 | 0.215 | Not significant |

**Table 4:**
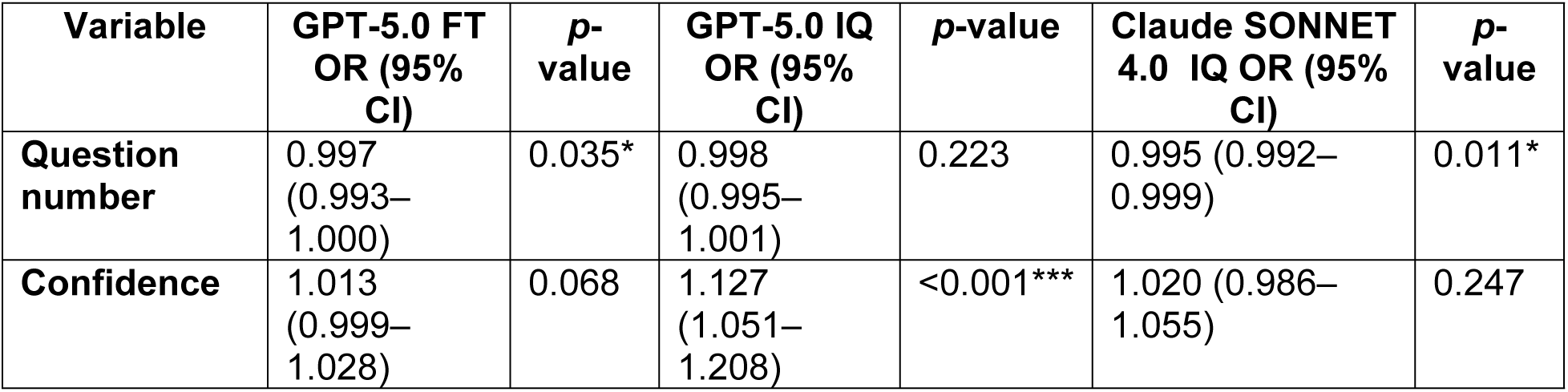
Logistic Regression Analysis of Question Order and Confidence as Predictors of Accuracy Across Models.

| Variable | GPT-5.0 FT<br>OR (95%<br>CI) | p-value | GPT-5.0 IQ<br>OR (95%<br>CI) | p-value | Claude SONNET<br>4.0 IQ OR (95%<br>CI) | p-value |
| --- | --- | --- | --- | --- | --- | --- |
| Question<br>number | 0.997<br>(0.993–<br>1.000) | 0.035* | 0.998<br>(0.995–<br>1.001) | 0.223 | 0.995 (0.992–<br>0.999) | 0.011* |
| Confidence | 1.013<br>(0.999–<br>1.028) | 0.068 | 1.127<br>(1.051–<br>1.208) | <0.001*** | 1.020 (0.986–<br>1.055) | 0.247 |

**Table 5:** Univariable Logistic Regression - Citation Validity Predicting Accuracy.

| Model | n | Citation OR (95% CI) | p-value | Percentage Effect |
| --- | --- | --- | --- | --- |
| GPT-5.0 FT | ~264 | **0.686** (0.370-1.272) | **0.232** | **-31.4%** |
| GPT-5.0 IQ | ~328 | **1.359** (0.376-4.909) | **0.640** | **+35.9%** |
| Claude SONNET 4.0 IQ | ~322 | **0.883** (0.475-1.642) | **0.694** | **-11.7%** |

Paired question-level analyses further supported this trend. McNemar’s test revealed that GPT-4.0 achieved more correct responses in IQ mode than FT mode on discordant items (39 vs. 23; χ² = 3.63, *p* = 0.057), indicating a performance improvement approaching statistical significance. A similar pattern was observed for GPT-5.0, though the difference was not statistically significant (38 vs. 27; χ² = 1.54, *p* = 0.215). Together, these results suggest that presenting questions individually may confer practical advantages by reducing contextual interference or cognitive overload, leading to modest but consistent performance gains.

### Calibration: Confidence–Accuracy Relationship

Binary logistic regression analysis demonstrated that confidence scores were statistically significant predictors of response accuracy across all models (*p* < 0.001) (Table X). GPT-5.0 in the Individual Question format showed the strongest association (OR = 1.125 per confidence point), indicating a 12.5% increase in the odds of a correct response for each unit increase in confidence. Claude SONNET 4.0 exhibited the highest overall classification accuracy (86.4%), although with the weakest discriminative ability (ROC AUC = 0.589), while GPT-5.0 FT demonstrated moderate predictive performance (OR = 1.018). All models showed excellent sensitivity (99.6–99.7%), reflecting reliable prediction of correct answers, but poor specificity (0–1.9%), indicating limited ability to identify incorrect responses. These findings confirm that model-generated confidence scores reliably tracked internal certainty and were significantly associated with accuracy, supporting their potential utility for evaluating AI reliability in clinical and educational settings.

### Model Accuracy Degradation Across token length

GPT-5.0 Full Test demonstrated significant question fatigue (OR = 0.997 per question, p = 0.035) with strong linear decline (R² = 0.784), indicating a 0.3% decrease in accuracy odds per question. Confidence showed marginal significance (OR = 1.013, p = 0.068). GPT-5.0 Individual Question mode showed no significant question fatigue effect (OR = 0.998, p = 0.223) with minimal linear correlation (R² = 0.095), but maintained strong confidence-accuracy calibration (OR = 1.127, p < 0.001). Claude SONNET 4.0 Individual Question exhibited the most pronounced question fatigue (OR = 0.995 per question, p = 0.011) with moderate linear decline (R² = 0.666) and no significant confidence effect (OR = 1.020, p = 0.247).

Scatter plots in figure 4 illustrate the relationship between question position (x-axis) and predicted probability of a correct response (y-axis) derived from multivariable logistic regression models for three large language models. Each panel represents a different model and evaluation format: (A) GPT-5.0 – Full Test condition, (B) GPT-5.0 – Individual Question condition, and (C) Claude SONNET 4.0 Individual Question condition. Blue points indicate predicted probabilities for individual questions, with solid lines representing fitted regression trends and shaded areas showing 95% confidence intervals. All three models exhibited a downward trajectory in accuracy as question number increased, suggesting potential cumulative context effects. This decline was most pronounced and statistically significant in GPT-5.0 Full Test mode (OR = 0.997 per question, *p* = 0.035, R² = 0.784) and in Claude SONNET 4.0 Individual Question mode (OR = 0.995, *p* = 0.011, R² = 0.666), while GPT-5.0 Individual Question showed a milder, non-significant trend (OR = 0.998, *p* = 0.223, R² = 0.095).

**Figure 4:**
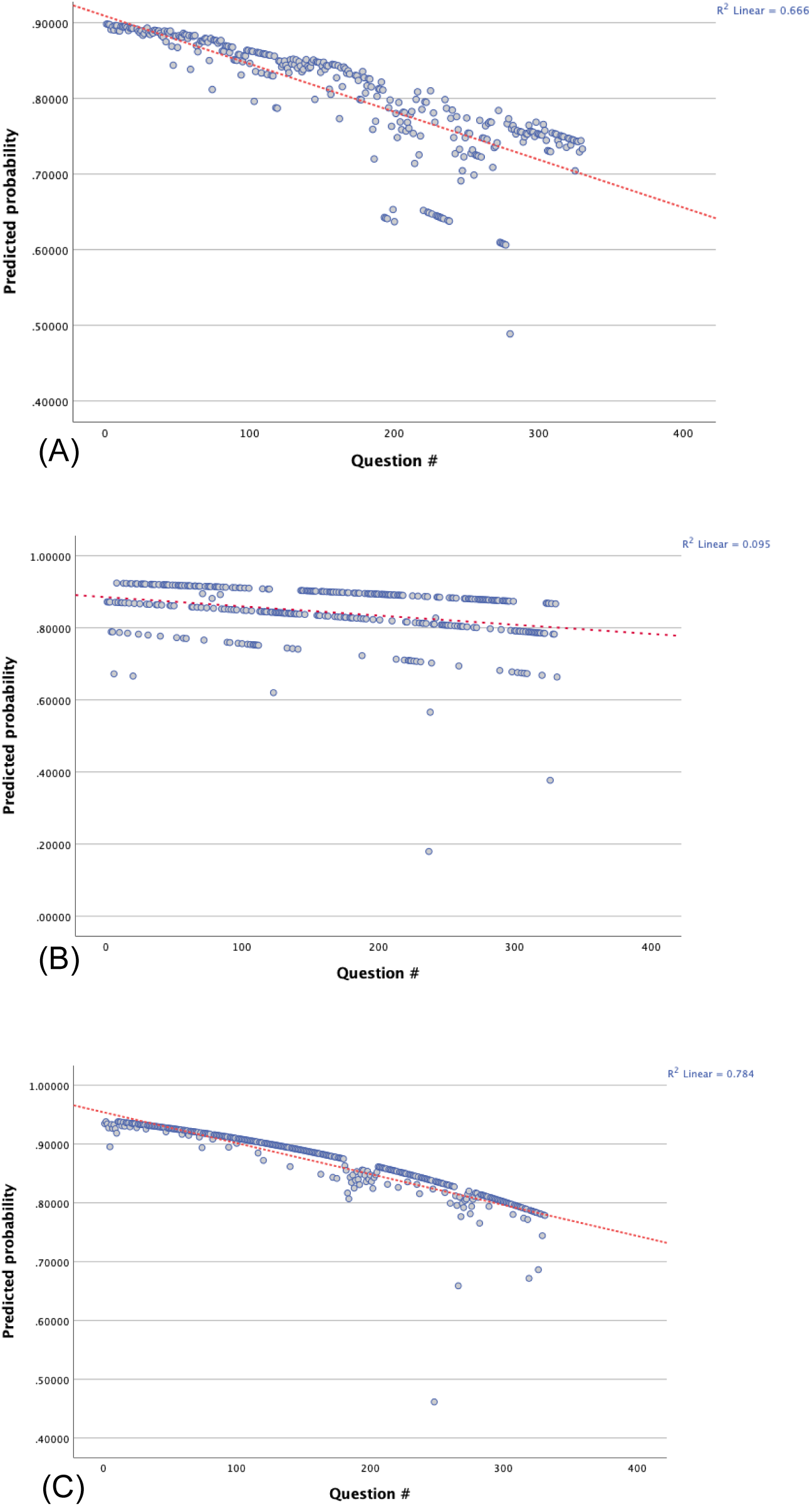
Multivariable logistic regression analyses examining the relationship between question position (proxy for cumulative context exposure) and model accuracy are shown for three large language models. Each panel plots the predicted probability of a correct response (y-axis) against question number (x-axis), with fitted regression lines and 95% confidence intervals. (A) GPT-5.0 in Full Test mode demonstrated significant question fatigue (OR = 0.997 per question, p = 0.035, R² = 0.784), indicating a progressive 0.3% reduction in accuracy odds per question. (B) GPT-5.0 in Individual Question mode showed no significant decline across question sequence (OR = 0.998, p = 0.223, R² = 0.095), maintaining stable performance with strong confidence–accuracy calibration (OR = 1.127, p < 0.001). (C) Claude SONNET 4.0 in Individual Question mode exhibited a significant decline in accuracy with increasing question number (OR = 0.995 per question, p = 0.011, R² = 0.666), consistent with model-specific susceptibility to cumulative context effects.

### Citation Validity and Accuracy

In contrast to confidence scores, citation validity did not significantly predict response accuracy for any model (*p* > 0.05) (Table 6). Although effect directions varied—with valid citations associated with a 35.9% increase in odds of a correct response for GPT-5.0 IQ and decreases of 31.4% and 11.7% for GPT-5.0 FT and Claude SONNET 4.0, respectively—none of these associations were statistically significant.

### Citation Hallucination Assessment

Citation validity evaluation revealed substantial differences across models. Claude SONNET 4.0 demonstrated the lowest hallucination rate at 5.59% (18/322), GPT-4.0 IQ showed 32.01% (105/328), and GPT-5.0 FT exhibited the highest rate at 51.53% (135/262). Citations were systematically classified using a three-tier validation system: fully verifiable, partially correct (minor errors in volume/page numbers), and critically wrong (fabricated authors, studies, journals, or completely unrelated content).

The heatmap in Figure 5 illustrates citation validity across three large language models (5.0 FT, IQ5.0 IQ, and Claude SONNET 4.0) for 331 questions organized into nine topical sections (S1–S9). Each thin vertical strip represents one question’s citation score, color-coded from red (0 = critically wrong) through orange (1 = partially correct, multiple errors) and yellow (2 = partially correct, fine errors) to green (3 = fully verified), with gray indicating missing responses. Claude SONNET 4.0 exhibits a predominance of green strips (>90% fully verified) in all sections, reflecting consistently high citation accuracy. IQ5.0 IQ shows substantial green interspersed with orange and red strips—approximately two-thirds fully verified—indicating moderate performance. In contrast, 5.0 FT displays frequent red and gray gaps, especially in sections S4, S7, and S8, underscoring its lower overall reliability and higher rates of missing or incorrect citations. The black vertical separators delineate topical sections, revealing that some sections (e.g., S5 and S6) pose greater challenges for all models, as evidenced by increased yellow and red densities. Overall, this compact visualization highlights clear performance gradients among the models and identifies specific content areas where citation validity deteriorates.

**Figure 5:**
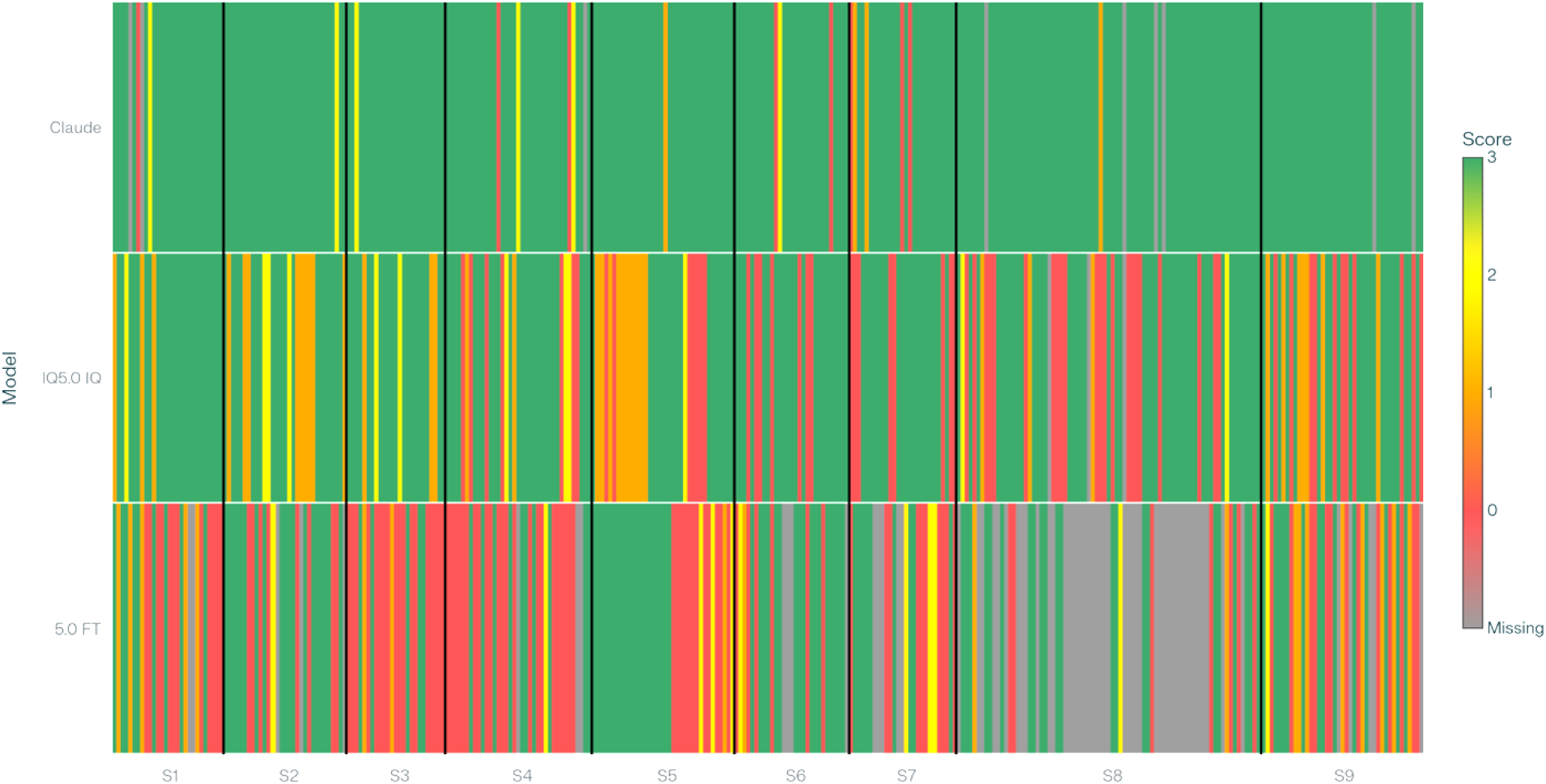
Heatmap showing citation validity across nine exam sections for three LLMs, with green indicating valid citations, yellow partially valid, red invalid, and gray missing.

Citation performance varied markedly by question category and model configuration (Figure 6). For non-specific factual questions, Claude SONNET 4.0 demonstrated the highest citation validity, with 96% of references fully verified, compared with 61% for GPT-5.0 IQ and 41% for GPT-5.0 FT. Notably, GPT-5.0 FT exhibited the highest incidence of critically incorrect citations (45%) in this category. Across author-specific questions, all models achieved improved citation accuracy, with Claude SONNET 4.0 verifying 93% of references, GPT-5.0 IQ 63%, and GPT-5.0 FT 61%, suggesting that contextual cues such as author names enhance citation reliability.

**Figure 6:**
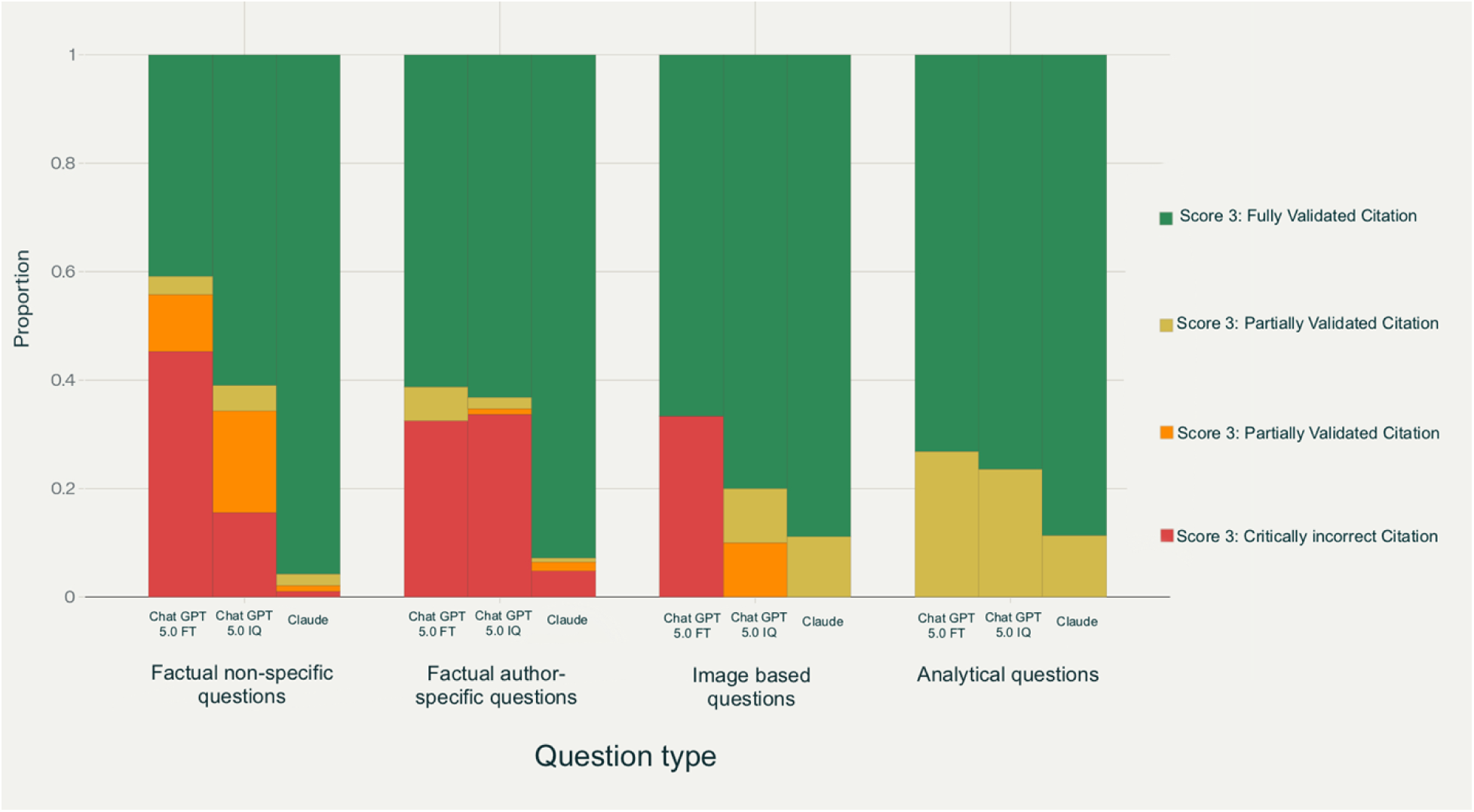
Proportion of citation validity outcomes by question type for three LLM configurations (GPT-5.0 FT, GPT-5.0 IQ, and Claude SONNET 4.0).

Image-based questions were also handled with relatively high validity, with Claude SONNET 4.0 verifying 89% of citations and GPT-5.0 IQ 80%, while GPT-5.0 FT achieved 67%. Analytical questions produced the most accurate outputs relative to non-specific factual items: Claude SONNET 4.0 verified 89% of citations, GPT-5.0 IQ 76%, and GPT-5.0 FT 73%, with most remaining errors classified as partially valid rather than critically incorrect.

### Relationship of Citation Source to Citation Validity

Pearson’s chi-square tests revealed a highly significant association between citation source type (article vs. book) and citation validity across all three models (χ² ≥ 236, df = 3, Bonferroni-adjusted p < 1 × 10⁻⁵⁰). These results indicate that the likelihood of a citation being verified is strongly dependent on the type of source from which it is drawn.

In practical terms, all models demonstrated a higher proportion of fully validated citations when referencing journal articles compared with books. For example, in the GPT-5.0 FT condition, approximately 62% of article-derived citations were fully verified compared with only 36% of book-based citations, with the latter contributing disproportionately to critically incorrect or missing references. Similar patterns were observed for GPT-5.0 IQ and Claude SONNET 4.0, underscoring a systematic bias in citation reliability based on source type.

## Discussion

Large language models (LLMs) have demonstrated substantial advancements in capability over recent years, with increasing integration across biomedical research, clinical practice, and professional education. (13)These models exhibit sophisticated capacity for synthesizing complex information and generating contextually appropriate responses, establishing their utility as computational tools within scientific and clinical frameworks.

Nevertheless, responsible implementation necessitates comprehensive evaluation methodologies that transcend conventional performance metrics.

Conventional assessment paradigms have predominantly employed static benchmarks including the Massive Multitask Language Understanding (MMLU), TruthfulQA, and General Language Understanding Evaluation (GLUE) tasks. (14) While these standardized assessments provide foundational performance data, they demonstrate limited capacity to predict model behavior under real-world valid conditions. Contemporary evaluation initiatives within biomedical domains have concentrated primarily on task accuracy, operationally defined as the proportion of correct responses under controlled experimental conditions. The present investigation, while not constituting a comprehensive system-level or longitudinal assessment, was designed within this methodological framework. The study focused on critical behavioral dimensions including response accuracy, confidence calibration, citation validity, and hallucination frequency, thereby addressing key considerations for clinical and educational applications.

### Comparing Model Performances

Our findings demonstrate that commercially available LLMs achieve substantial performance levels on specialty-specific assessments, with overall accuracy ranging from approximately 78% to 87% depending on model architecture and delivery methodology. These results corroborate emerging literature documenting progressive performance improvements across successive LLM generations and establish that general-purpose models, absent domain-specific fine-tuning, can effectively respond to complex professional-level inquiries. However, accuracy exhibited heterogeneous distribution across content domains.

Superior performance was observed in knowledge-intensive domains including biochemistry, physiology, and microbiology, where questions predominantly require retrieval of well-established factual information. Conversely, integrative domains—particularly diagnosis and periodontal therapy—presented greater challenges across all evaluated models.

Performance analysis across question typologies revealed that LLMs demonstrated proficiency with both factual and analytical question formats. Notably, contextualized questions yielded superior performance compared to broader conceptual inquiries, as illustrated in the radar plot visualization. The observed superiority of LLM performance on factual versus analytical periodontal questions aligns with patterns documented in a comprehensive systematic review and network meta-analysis by Wang et al. Their analysis of 35,896 medical questions across 168 studies demonstrated consistent LLM superiority on “objective questions” (characterized by clear, quantifiable responses) compared to “open-ended questions” (requiring complex reasoning without predetermined solutions).(6) This performance pattern remained consistent across examination sections despite variations in model training methodologies. Both full-test and individual question implementations of ChatGPT 4.0, ChatGPT 5.0, and Claude SONNET 4.0 sonnet 4.0 exhibited comparable relative performance hierarchies across subject domains. Network visualization analysis revealed that while individual models demonstrated varying overall performance, subject difficulty rankings remained remarkably stable, indicating that specific dental education domains present inherent challenges for current LLM architectures. Cross-generational model performance analysis revealed that GPT-5.0 demonstrated modest yet consistent improvements relative to GPT-4.0 across all testing conditions. The most substantial performance gain was observed in the full-test format (+2.74 percentage points), with attenuated differences in the individual-question format (+1.51 percentage points). However, these differences did not achieve statistical significance (χ² = 0.850, p = 0.357; Cramer’s V = 0.025), indicating that while iterative model refinements contribute to incremental accuracy improvements, their impact remains constrained within specialized domain assessments.

These findings present a notable contrast to broader benchmark evaluations that document substantial performance gains for GPT-5 relative to GPT-4 across various domains. In standardized assessments, GPT-5 achieved 94.6% accuracy on the AIME 2025 mathematics benchmark compared to GPT-4’s approximately 52% performance and attained 74.9% accuracy on SWE-bench Verified coding tasks versus GPT-4’s 30-50% range. (15,16)

Healthcare applications demonstrate a more nuanced performance profile. While GPT-5 exhibited reduced hallucination rates to 1.6% on selected medical benchmarks compared to GPT-4o’s 15.8%, performance varies considerably across different healthcare tasks, with heterogeneous results on clinical reasoning and medical knowledge assessments.(16,17)The observed disparity between general benchmark improvements and domain-specific performance suggests that while GPT-5 demonstrates marked advances in structured cognitive tasks such as mathematics and coding, its impact on specialized professional applications remains inconsistent. (18) This pattern indicates that generational improvements in large language models may not uniformly translate to enhanced performance across all professional domains, necessitating domain-specific evaluation frameworks for clinical and educational applications.

### Comparing the method of data presentation

LLMs are rapidly evolving, and the ways in which users interact with them continue to adapt as our understanding of their capabilities deepens. Prompt engineering has always been central to optimizing model performance; however, our approach to it has matured — moving from the assumption that longer, more elaborate prompts are inherently better to the recognition that prompts should instead be information-rich, detailed, and contextually specific, yet concise and focused.(19) Another critical dimension of LLM behavior relates to the influence of input length and conversational duration on accuracy.

Several studies demonstrate that performance can degrade as context length increases, with models showing diminished ability to retrieve or reason over information buried deeper within extended inputs. This so-called “lost-in-the-middle” phenomenon underscores how the structure and sequencing of information directly affect output quality. (20,21) Some models, such as Claude SONNET 4.0, even prompt users to begin a new conversation when the input becomes too long, reflecting the practical implications of this limitation.

To examine how these dynamics shape model performance, our study deliberately varied data presentation strategies: in one condition, the entire assessment was provided at once to maximize contextual availability, whereas in another, questions were presented sequentially to minimize token load. This design allowed us to investigate whether well-documented declines in performance with increasing context length persist across different presentation modes and whether strategic segmentation can mitigate accuracy loss in specialized tasks.

Our results reveal two distinct but complementary phenomena. First, presentation format demonstrated a consistent directional effect on outcome accuracy. Individual Question presentation produced improvements across both models, with combined analysis approaching statistical significance (χ² = 3.41, p = 0.065). GPT-4.0 showed a 4.86 percentage point improvement (p = 0.140), while GPT-5.0 demonstrated a 3.34 percentage point gain (p = 0.305). McNemar’s paired analysis corroborated this pattern, with GPT-4.0 approaching significance (p = 0.057) and GPT-5.0 showing similar directional effects (p = 0.215). Although statistical significance was not achieved, the directional consistency across models suggests that individual question presentation may mitigate contextual interference or cognitive overload, leading to modest but practically relevant performance improvements.

Second, analysis of question order as a proxy for increasing token load revealed differential’question fatigue’ effects across models and presentation formats. GPT-5.0 in full-test mode demonstrated significant accuracy degradation, with odds of a correct response decreasing by 0.3% per question (OR = 0.997, p = 0.035, R² = 0.784). This effect was not observed in GPT-5.0 individual question mode, which showed no significant decline (OR = 0.998, p = 0.223, R² = 0.095). Claude SONNET 4.0 in individual question mode also exhibited significant question fatigue (OR = 0.995, p = 0.011, R² = 0.666), with a more pronounced decline in accuracy across the sequence, although its overall accuracy remained high. These findings suggest that question fatigue is not simply a function of cumulative computational burden, but rather reflects complex interactions between model architecture, presentation format, and context management. The progressive decline in full-test mode aligns with theories of attention saturation and positional encoding inefficiency in transformer architectures, consistent with Levy et al. (2024) findings of decreased reasoning accuracy as input length increased from 250 to 3000 tokens.(22)

### Confidence and Calibration: Insights into Model Self-Assessment

Confidence estimation was incorporated as a key parameter because it reflects an AI model’s capacity to assess the reliability of its own outputs, a critical property for effective deployment in educational and clinical decision-support contexts. Optimal performance requires that self-reported confidence align closely with the probability of correct responses, a characteristic termed calibration. Analysis revealed that while confidence estimates from both GPT-5.0 and Claude SONNET 4.0 were not perfectly calibrated to response accuracy, they demonstrated statistically significant predictive relationships across all evaluated systems.

Binary logistic regression confirmed that confidence scores served as significant predictors of model accuracy, with GPT-5.0 in Individual Question mode exhibiting the strongest predictive association, followed by Claude SONNET 4.0 and GPT-5.0 in Full Test mode. This hierarchy suggests that segmented input presentation enhances confidence-accuracy calibration.

The observed relationship between confidence and accuracy, while statistically significant, indicates that confidence metrics provide useful but imperfect indicators of response reliability. These findings underscore the necessity for complementary validation mechanisms capable of more effectively identifying uncertain or potentially incorrect responses in high-performing AI systems deployed in professional contexts.

### Citation Integrity and Hallucination

Perhaps the most consequential limitation revealed by this study was the substantial variability in citation quality. While both GPT-5.0 and Claude SONNET 4.0 generated references when prompted, a considerable proportion were partially incorrect or wholly fabricated—a phenomenon termed “citation hallucination.” This behavior reflects the generative nature of LLMs, which predict text patterns rather than retrieve verified database records. Although models frequently produced references with plausible formatting and content, many could not be traced to authentic sources or failed to support their associated claims. This limitation is particularly problematic in educational and research settings, where users may rely on citations for further reading, evidence validation, or scholarly writing. Fabricated references not only undermine user trust but also risk propagating misinformation if accepted uncritically.

Analysis revealed that citation validity was independent of answer accuracy, similar to seen by Danesh et al, 2025. (23) Human-in-the-loop assessment demonstrated that hallucinated citations existed across a spectrum of severity. The three-tier classification system differentiated between fully verifiable citations, partially correct citations with minor errors (incorrect volume numbers, page references), and critically erroneous citations containing fabricated authors, non-existent studies, incorrect journal attributions, or wholly unrelated content. The predominance of critically erroneous hallucinations—rather than minor bibliographic inaccuracies—indicates that AI citation fabrication represents a qualitatively distinct problem from human citation errors, consistent with findings reported by Aljamaan et al. (24)

Citation performance varied systematically across question categories. Author-specific and analytical questions achieved consistently higher citation accuracy across all models compared to non-specific factual questions, suggesting that contextual specificity significantly influences citation reliability. Non-specific factual questions exhibited the highest incidence of critically incorrect citations, paralleling patterns observed in answer accuracy metrics. These findings underscore the critical need for enhanced citation validation frameworks, including improved retrieval-augmented generation (RAG) mechanisms, real-time database cross-referencing systems, or hybrid architectures that integrate generative capabilities with verified bibliographic databases. Such implementations would ensure that model-generated references maintain both authenticity and contextual relevance, thereby addressing the fundamental disconnect between pattern-based text generation and evidence-based scholarly citation practices.

### Broader Implications and Future Directions

These findings collectively demonstrate both the potential and current constraints of LLMs as educational and knowledge-support tools in specialized dental practice. The observed high overall accuracy indicates that such models can effectively augment learning processes and assessment preparation, particularly for reinforcing foundational knowledge and simulating examination-style questioning protocols. However, persistent limitations in reasoning-intensive clinical domains, suboptimal confidence calibration, and systematic citation reliability deficits necessitate continued human oversight for safe and effective implementation.

Future investigations should extend this multidimensional evaluation framework through several key directions: (1) longitudinal assessments spanning multiple examination cycles to evaluate temporal stability of model performance, (2) expansion of citation validation protocols incorporating larger, diverse reviewer panels to enhance reliability of bibliographic assessment, and (3) integration of advanced calibration metrics including Brier scores, expected calibration error, and reliability diagrams to provide more nuanced confidence-accuracy relationships.

Additionally, comparative analyses across multiple dental specialties and integration of real-world clinical scenarios would strengthen the ecological validity of these findings. As LLM architectures continue advancing, such comprehensive, specialty-specific evaluation protocols will prove essential for guiding evidence-based integration into dental education curricula, clinical decision-support systems, and scholarly research workflows. The framework established in this study provides a foundation for systematic assessment of emerging AI technologies in professional healthcare education contexts.

## Conclusion

While large language models demonstrate the capacity to answer a wide range of dental assessment questions, their reliability is variable and highly dependent on both the context and the manner in which information is presented. This variability underscores that, although LLMs can serve as valuable adjuncts for knowledge reinforcement and assessment preparation, their outputs cannot be assumed accurate or trustworthy in all scenarios. In educational and research applications, it is essential that all LLM-generated responses, particularly those involving complex reasoning or citation are subject to rigorous human review. Ongoing human oversight and validation remain critical to ensure the integrity and safety of information, and future research should focus on developing robust frameworks for integrating LLMs responsibly within dental education and scholarly practice.

## Ethics Approval Statement

This study did not involve human participants or identifiable patient data. All questions were derived from publicly available educational resources. The study was conducted in alignment with principles of responsible AI research, emphasizing transparency, reproducibility, and human oversight in citation validation.

## Author Contributions

P.D. conceived the study design, conducted data collection and assessment, performed statistical analysis, and drafted the manuscript. R.H. contributed to study conception and data collection. Both authors reviewed and approved the final version of the manuscript.

## Data Availability

The datasets generated and analyzed during this study are available from the corresponding author upon reasonable request. The complete dataset includes: AI model responses to all 331 American Academy of Periodontology examination questions, including accuracy assessments, confidence scores, and generated citations for GPT-4o, GPT-4o mini, and Claude-3.5 Sonnet models Human expert validation results for all AI-generated citations, including verification status and authenticity scores Statistical analysis outputs including confidence calibration metrics, accuracy measurements across question categories, and question fatigue analysis. Data requests should be directed to the corresponding author

## Acknowledgements

The authors thank Hui Bian, Ph.D., Research & Statistics Consultant, Office for Faculty Excellence, East Carolina University, for providing valuable statistical consultation and guidance during the data analysis phase of this study.

